# Antibiotic-free vaginal microbiota transplantation (VMT) changes vaginal microbiota and immune profile in women with asymptomatic dysbiosis – reporting of a randomized, placebo-controlled trial

**DOI:** 10.1101/2024.06.25.24309408

**Authors:** Elleke F. Bosma, Brynjulf Mortensen, Kevin DeLong, Mads A. Røpke, Helene Baek Juel, Randi Rich, Amalie M. Axelsen, Marouschka J. Scheeper, Rasmus L. Marvig, Thomas Gundelund Rasmussen, Colleen Acosta, Ulrich K. Binné, Anne Bloch Thomsen, Hans-Christian Ingerslev, Fareeha Zulfiqar, Tine Wrønding, Paul D. Cotter, Marcus O’Brien, Shriram Patel, Sarita A. Dam, Julia Albert Nicholson, Henriette Svarre Nielsen, Timothy G. Dinan, Fergus P. McCarthy, Johan E.T. van Hylckama Vlieg, Laura M. Ensign

## Abstract

Here, we describe the first placebo-controlled trial of vaginal microbiota transplantation (VMT) in women with asymptomatic dysbiosis without the use of antibiotic pretreatment. Importantly, we also describe the implementation of a donor program and banking of donor cervicovaginal secretions (CVS) while retaining sample viability, which is crucial to allow for scale-up and confirmatory quality testing. By metagenome sequencing, we demonstrate that VMT provided a significant increase in combined *Lactobacillus* species in the active arm and strain-level genetic analysis confirmed *Lactobacillus* engraftment. Moreover, VMT was well tolerated and showed a good safety profile. Furthermore, a shift toward increased *Lactobacillus* was associated with a change in the expression profile of genes in the complement pathway to a more anti-inflammatory profile. Vaginal microbial and immune profile restoration using VMT may have a positive impact on a wide range of conditions in women’s health.

## Introduction

The composition of the vaginal microbiota has a profound impact on sexual and reproductive health^1-4^. The cervicovaginal tract contains commensal bacteria communities, wherein dominance by a subset of *Lactobacillus* species is generally associated with better health outcomes. A deviation from a low diversity community is often characterized by a high abundance of facultative and strictly anaerobic species and is regarded as a non-optimal state referred to as dysbiosis, that may or may not be accompanied by clinical symptoms^5^. This is in dramatic contrast to the more widely studied intestinal microbiota, where a loss of diversity is often observed at the onset or during disease, and a high bacterial diversity is key to maintenance of homeostasis^6^. The composition of the vaginal microbiota correlates with risk of infection of the reproductive and urinary tracts^5,7-10^, fertility and pregnancy outcomes^11-17^, and potentially cancers and cancer treatment outcomes^18-20^. However, despite the tremendous importance of the vaginal microbiota, we currently lack effective strategies for correcting dysbiosis durably.

“Symptomatic” dysbiosis of the vaginal microbiota is a condition called bacterial vaginosis (BV), which affects 29-52% of women depending on the study population and geographic location^21^. BV is not typically associated with symptoms such as redness, itching, or pain, which is why it is referred to as “vaginosis” rather than “vaginitis”^5^. When dysbiosis is accompanied by at least three out of four Amsel’s criteria, which include pH > 4.5, the presence of “clue” cells, discharge, and fishy odor, it is considered symptomatic BV. However, these diagnostic criteria are heavily dependent on the perception of the clinician and the patient, calling into question the diagnostic utility^5^. Regardless, the current clinical guideline is that only women with these clinical symptoms of BV would receive antibiotic treatment, and recurrence rates are as high as 70%^22,23^. Further, as high as 84% of women with vaginal dysbiosis identified by molecular techniques report no symptoms and are considered “asymptomatic”^24,25^. Importantly, dysbiosis has been shown to result in a more pro-inflammatory environment, regardless of whether clinical symptoms are present, and this is associated with the increased risk of infections and other negative health outcomes ^26-30^. Thus, regardless of whether symptoms are present or whether treatment is administered, the persistence and recurrence of dysbiosis and the related local inflammation is a major unaddressed issue in women’s health.

Due to the persistence and recurrence of vaginal dysbiosis, a wide variety of alternative experimental approaches are used with limited treatment success, including long-term intermittent antibiotic use, chemical cleansing agents, and probiotics^31,32^. One potentially more promising approach was inspired by the success of fecal microbiota transplant (FMT) in treating severe, recurrent *Clostridium difficile* infection and inflammatory bowel disease^33^. The premise of vaginal microbiota transplant (VMT) is also motivated by the concept that transplanting the whole microbiota community may be necessary to establish dominance by *Lactobacillus* species in the reproductive tract. Indeed, the first exploratory study testing the use of VMT as a therapeutic alternative for patients suffering from symptomatic, intractable, and recurrent BV showed great promise in a cohort of five patients^34^. One to three instances of antibiotic treatment followed by VMT with material from one or two unique donors resulted in long-lasting improvements in symptoms and molecular signs of BV in four of the patients^34^. While this study showed the potential of VMT as treatment, it used fresh donor material without storage prior to use, and it did not report strain-level engraftment of donor material. The ability to store donor material without loss of viability is crucial for scale-up and safety testing. Recently, our teams showed strain-level engraftment of banked donor material after antibiotic-free VMT in a patient with years of BV symptoms and recurrent pregnancy losses ^35^.

Here, we describe a placebo-controlled trial of antibiotic-free VMT using screened, frozen, and banked donor cervicovaginal secretion (CVS) material used to treat participants with asymptomatic dysbiosis (**Figure 1**). In a companion study by Wrønding et al., a similar donor program was applied to a recipient population that included subjects with symptomatic dysbiosis, and a different dosing regime was tested. Here, we demonstrate a significant increase in the relative abundance of vaginal lactobacilli following VMT, along with evidence of sustained engraftment of *L. crispatus* strains from the donor CVS in the VMT recipients. Importantly, when there was a dramatic shift in microbiome composition after the intervention, there was also a shift in expression of a range of genes in the complement pathway towards a less inflammatory state that was consistent with *Lactobacillus* dominance in a cross-sectional cohort. Our study shows that VMT has great potential as the first therapeutic intervention that provides effective and long-term vaginal microbiota restoration, positively impacting a wide variety of diseases and conditions affecting the female reproductive tract.

**Figure 1.**
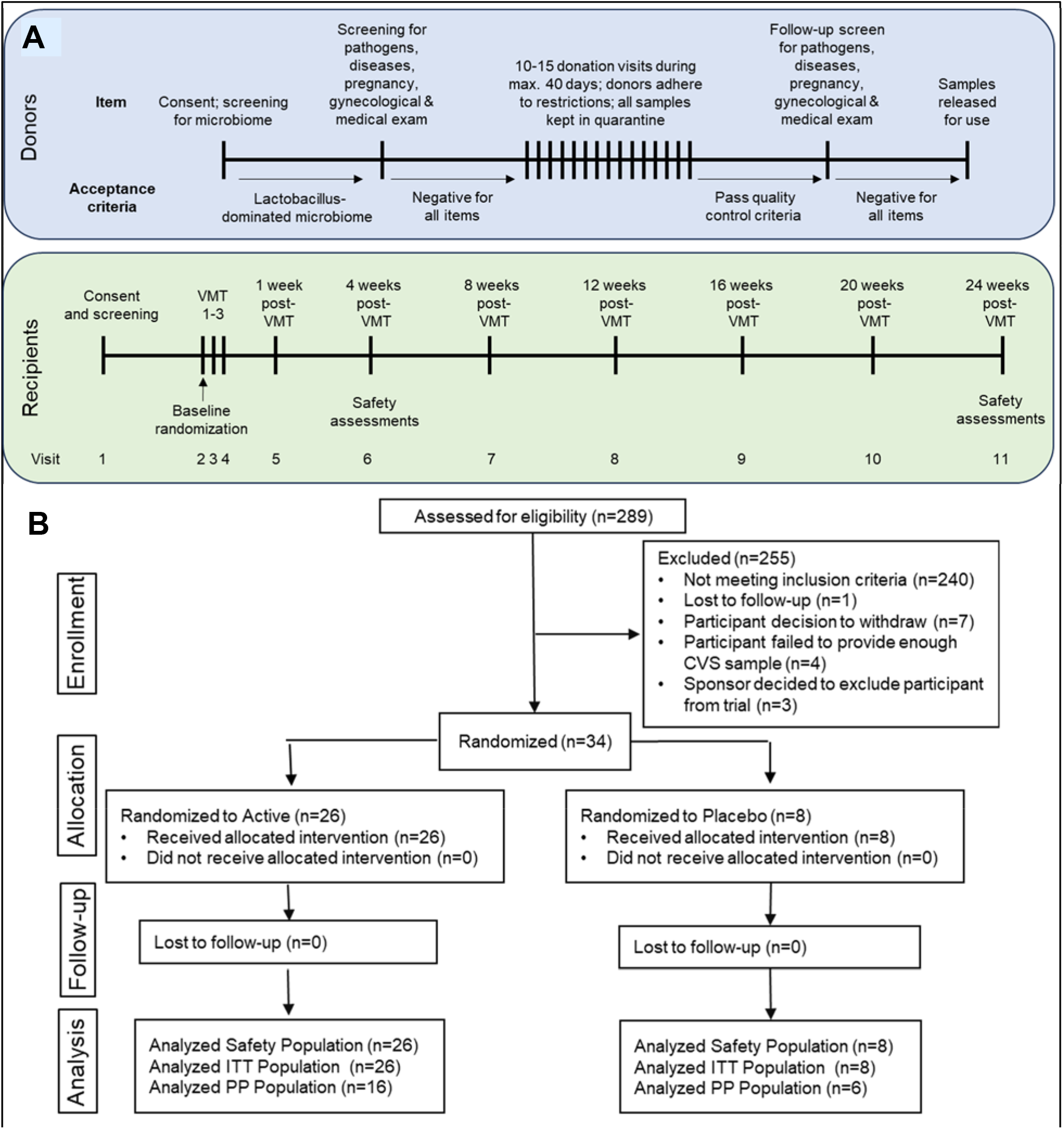
(A) Schematic representation of the flow of participants through the donor screening program. Potential participants were first screened for microbiota composition and tested for a wide range of infections and conditions. After passing the screening tests, subjects provided 10-15 CVS samples over a period of up to 40 days. The CVS samples were frozen and quarantined until the participant was again tested to confirm suitability of the samples for use in the VMT intervention. Samples were then characterized for potency and released for use. (B) Schematic representation of the flow of recipient participants through the VMT intervention study. Potential participants were first screened for microbiota composition and tested for a wide range of infections and conditions (Visit 1). If they passed the screening tests, they were randomized 3:1 to either the VMT or placebo intervention groups. Both groups received three consecutive dosing procedures with either donor CVS or placebo, spaced one day apart (Visits 2-4). Follow-up visits were held at 1 week (Visit 5) and 1 month (Visit 6) after VMT, with monthly visits through 6 months (Visits 7-11). (C) Diagram showing the flow of recipient participants in the intervention study. A total of 289 women were assessed for eligibility, and 34 were randomized 3:1 into the VMT (n = 26) and placebo arms (n = 8). No participants were lost to follow-up in either group. All participants in the intention-to-treat (ITT) group were also included in the safety assessments. A subset of the participants were included in the per-protocol (PP) group.

## Results

### Screening cohort to identify potential CVS donors

For the purposes of donor screening, an “optimal” vaginal microbiome was defined as >80% combined relative abundance of vaginal *Lactobacillus* spp. with <5% combined relative abundance of *Atopobium spp*., *Prevotella spp*., *G. vaginalis, and F. vaginae*. Of the 96 women screened, 61/96 (64%) fulfilled the “optimal” criteria **(Supplementary Figure 1A)**. In the majority (41/61, 67%), *L. crispatus* comprised >80% by relative abundance, and *L. iners* comprised >80% relative abundance in 7/61 (11%) of cases (**Supplementary Figure 1B**). The remainder comprised mixed *Lactobacillus* populations containing two or more of *L. crispatus, L. iners, L. jensenii*, and *L. gasseri* (**Supplementary Figure 1B**). A “dysbiotic” vaginal microbiome was defined as <10% combined relative abundance of vaginal *Lactobacillus* spp. with >20% combined relative abundance of *Atopobium spp*., *Prevotella spp*., *G. vaginalis, and F. vaginae*, which was found to occur in 27/96 (28%) of women, with 8/96 (8%) falling in neither the optimal nor the dysbiotic category (“undefined”) **(Supplementary Figure 1A)**. The vaginal microbiota of most women in the dysbiotic category was found to be dominated by *G. vaginalis*, and a mixture of G. *vaginalis* and *Lactobacillus* spp. was observed in most cases in the undefined category (**Supplementary Figure 1B**).

### CVS donor selection

Of the 61 women that were categorized as having optimal vaginal microbiota, 38 passed HPV screening (62%). Upon further screening, 26 of the women failed for reasons such as testing positive for HPV or other potential pathogens on follow-up or abnormal gynecological findings (**Supplementary Table 1**). Of the 12 women that passed all additional screening, 3 failed during the donation process or at the follow-up testing, leaving 9 women that provided CVS samples approved for the transplantation procedure (**Supplementary Table 1**). The bacterial relative abundances in the vaginal microbiota of all potential donors screened is shown in **Supplementary Table 2**. The demographic information collected for the 9 confirmed donors is shown in **Supplementary Table 3**. Each donor provided 10-15 CVS samples during the donation period (≤40 days), and several donors provided CVS samples during two donation periods with a 1-2 month gap between donation periods. CVS samples from 5 donors were consistently dominated by *L. crispatus*, CVS from 1 donor was consistently dominated by *L. iners*, and the CVS from the other donors contained a majority of two or more of *L. crispatus, L. iners*, and *L. jensenii* with fluctuations in relative abundance of each species **(Figure 2A**). **Supplementary Table 3** shows the range and mean values for sample mass, pH, and viable bacterial cells per CVS sample provided by each donor.

**Figure 2.**
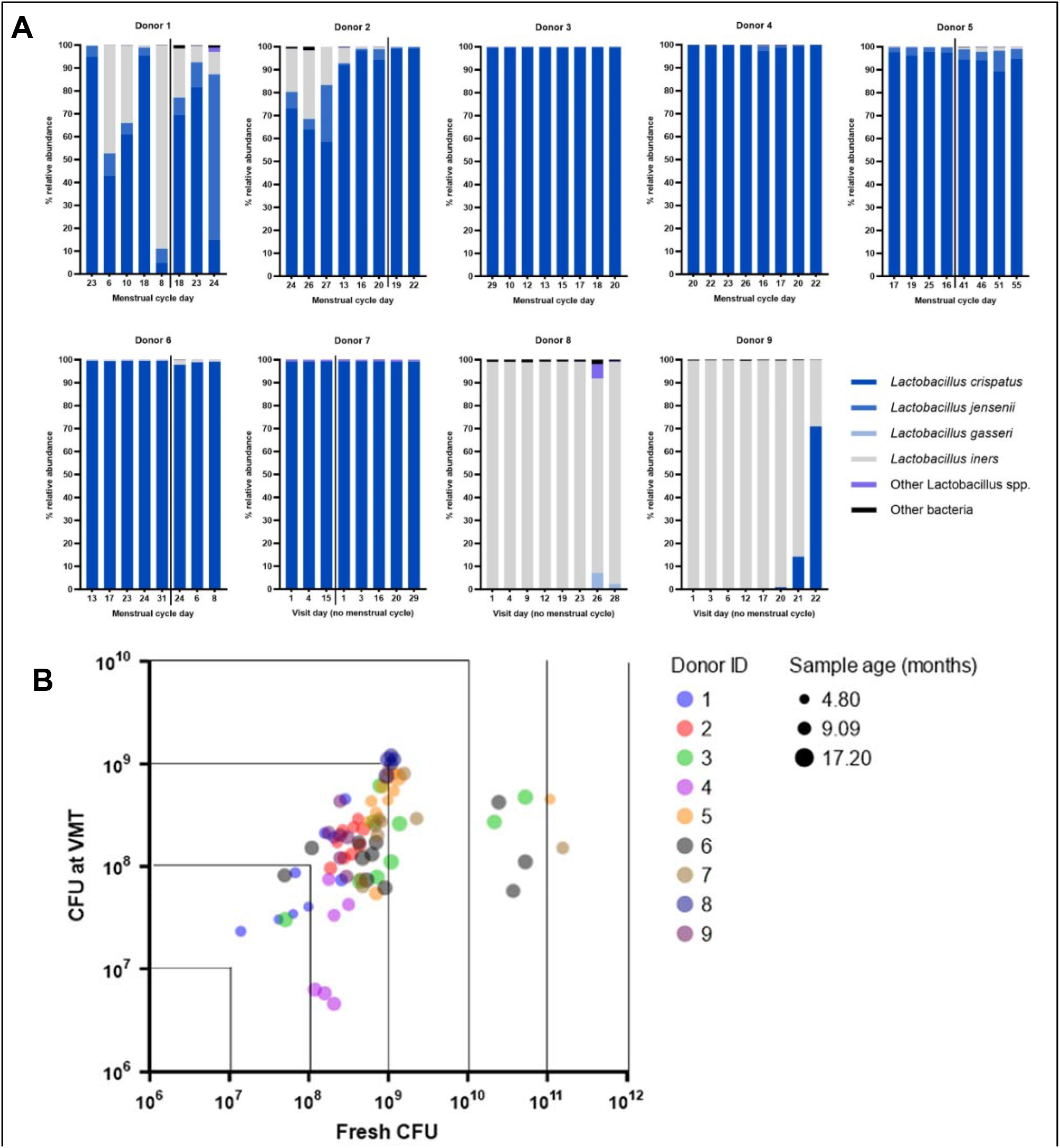
(A) Relative abundance of vaginal microbiota observed in donor CVS samples as characterized by sequencing. The relative abundance of the Lactobacillus species is shown for each CVS sample along with the menstrual cycle day collected. For each donor, representative selected visits are shown. For donors using contraceptives that suppress menstruation, the timing is noted as visit days to show the timing between samples. Vertical lines indicate a break in sample collection of 1-2 months, after which the donor was re-enrolled to provide additional samples in a subsequent donation period. (B) Relationship between the measured Lactobacillus colony forming units (CFU) in donor CVS samples at the time of collection (Fresh) compared to at the time of thawing and use for VMT. Each circle represents an individual sample used for VMT with color coding based on Donor ID. The size of the circle reflects the duration of freezer storage (sample age).

### Donor CVS freezer stability and quality control

For safety purposes, all donor CVS samples were frozen and quarantined until confirmatory donor testing (**Figure 1A**). CVS samples generally showed a reduction in bacterial cell viability after freezing, though there was no clear trend in terms of loss in potency during freezer storage (**Figure 2B**). When characterizing the bacterial cell count in the fresh CVS samples compared to when the samples were thawed for VMT, sample clustering highlighted that the loss in viability largely occurred during the freezing process rather than the storage process (**Figure 2B**). However, higher loss in bacterial cell count (2-3 logs) was observed in CVS samples with fresh bacterial cell counts >10E+10, though bacterial cell counts in all thawed donor CVS samples remained >10E+6 (**Figure 2B**).

### Screening and enrollment of VMT recipients

A total of 289 women were screened for eligibility to participate in the study as VMT recipients. Due to a higher than anticipated screening failure rate and COVID-related logistical challenges, 34 out of the planned 40 participants were randomized (26 Active, 8 Placebo) (**Figure 1B**). No participants in either group were lost to follow-up, so both the intention-to-treat (ITT) and the safety populations consisted of the 34 recipient participants. Twelve participants (10 Active, 2 Placebo) were excluded from the per-protocol (PP) analysis due to use of oral antibiotics or anti-fungal medications for reasons unrelated to the intervention during the study follow-up period (**Figure 1B**). The recipient participants in the Active and Placebo arms (both ITT and PP) were similar in terms of age, body mass index, ethnicity, smoking status, alcohol consumption, and use of hormonal contraception (**Table 1**).

**Table 1.** Demographics of the recipient participants that were included in the intention-to-treat (ITT) and the per protocol (PP) populations. All recipient participants that were excluded from the PP analysis used either oral antibiotics or anti-fungals (10 in the active group and 2 in the placebo), the use of which was excluded in the protocol. Data shown as mean with SD.

|  | ITT (n=34) |  |  |  | PP (n=22) |  |  |  |
| --- | --- | --- | --- | --- | --- | --- | --- | --- |
|  | Active (n=26) |  | Placebo (n=8) |  | Active (n=16) |  | Placebo (n=6) |  |
|  | Mean (SD) |  | Mean (SD) |  | Mean (SD) |  | Mean (SD) |  |
| Age (years) | 30.62 (9.26) |  | 31.13 (7.94) |  | 32.13 (9.43) |  | 28.67 (7.53) |  |
| BMI (kg/m<sup>2</sup>) | 26.11 (6.00) |  | 28.12 (7.21) |  | 25.91 (6.74) |  | 25.26 (5.27) |  |
| **Ethnicity** | **n** | **%** | **n** | **%** | **n** | **%** | **n** | **%** |
| White - Irish | 20 | 76.9 | 7 | 87.6 | 13 | 81.3 | 5 | 83.3 |
| White – Any other White background | 5 | 19.2 | 1 | 12.5 | 2 | 12.5 | 1 | 16.7 |
| Other (Irish/Lebanese) | 1 | 3.8 | - | - | 1 | 6.3 | - | - |
| **Other characteristics** | **n** | **%** | **n** | **%** | **n** | **%** | **n** | **%** |
| Current Smoker | 6 | 23.1 | 3 | 37.5 | 4 | 25.0 | 1 | 16.7 |
| Consume alcohol | 20 | 76.9 | 5 | 62.5 | 13 | 81.3 | 4 | 66.7 |
| Hormonal Contraceptive User | 10 | 38.5 | 3 | 37.5 | 6 | 37.5 | 3 | 50.0 |

### Recipient safety analyses

All vital signs were within the normal ranges at screening and baseline, and there were no significant changes in the median values for all vital signs at weeks 4 and 24 in the Active and Placebo groups (not shown). All recipient participants were tested for a range of possible infections at 4 weeks after the VMT procedure, and there were no incident infections attributable to the VMT procedure (**Supplementary Table 4**). All clinical chemistry and hematology parameters were in the normal ranges at baseline, and there were no significant differences in median values at 4 or 24 weeks after the procedure in both the Active and Placebo groups (not shown). A few subjects in both the Active (**Supplementary Table 5**) and Placebo (**Supplementary Table 6**) groups had high or low clinical chemistry or hematology parameters, leading to the recording of adverse events (AEs) that were not deemed clinically significant or related to the intervention. Ninety-five AEs were recorded, including 76 in the Active group (**Supplementary Table 5**) and 19 in the Placebo group (**Supplementary Table 6**). Thirty-two additional AEs were classified as adverse events of special interest (AESIs) (25 Active, 7 Placebo) (**Supplementary Table 7**). There were 23 AESIs in the Active group and 4 in the Placebo group that were deemed related to the intervention, including changes in menstrual cycle and intermenstrual bleeding (**Supplementary Table 7**). No serious AEs (SAEs) were recorded, and none of the AEs led to early termination of participation in the study (not shown).

### *Lactobacillus* composition after VMT

The primary objective was the change in overall vaginal lactobacilli (combination of *L. crispatus, L. iners, L. gasseri*, and *L. jensenii)* by relative abundance. There was an increase in the combined relative abundance of vaginal lactobacilli after VMT compared to Placebo in both the ITT (**Figure 3A**) and PP groups (**Figure 3B**). In the PP group, the combined relative abundance of lactobacilli compared to baseline was significantly increased up to Week 8 after VMT and showed a persistent trend for improvement out to 24 weeks, while there was no significant difference in the Placebo group compared to baseline at any follow-up visit (**Table 2**). Interestingly, the measured CFU at the time of dosing, whether compared to the fresh CFU or the duration of freezer storage, was not predictive of whether the donor CVS resulted in an increase in lactobacilli in the recipient (not shown). If *L. iners* was excluded from the analysis, then there was a clearer difference between the mean combined relative abundance in the Active group compared to Placebo in both the ITT (**Figure 3C**) and PP groups (**Figure 3D**). However, the mean values did not capture the bimodal distribution of *Lactobacillus* prevalence, which is further highlighted by visualizing the bacterial relative abundance over time in each recipient (**Figure 3E**). To differentiate between the expansion of endogenous *Lactobacillus* and the engraftment of donor strains in recipients, single nucleotide variant (SNV) analysis was performed. This was performed for all recipients in the active arm, of which four representative examples numbered 1-4 are shown in **Figure 3E**, were used in the analysis. For subjects 1 and 2, the *L. crispatus* observed after VMT was shown to be more genetically similar to the strains in the matched donor CVS samples than the CVS samples from all other donors (**Figure 3F**). In contrast, for recipients 3 and 4 that had little detectable *L. crispatus* after VMT, any strains detected were genetically dissimilar to all donor strains (**Figure 3F**).

**Table 2.**
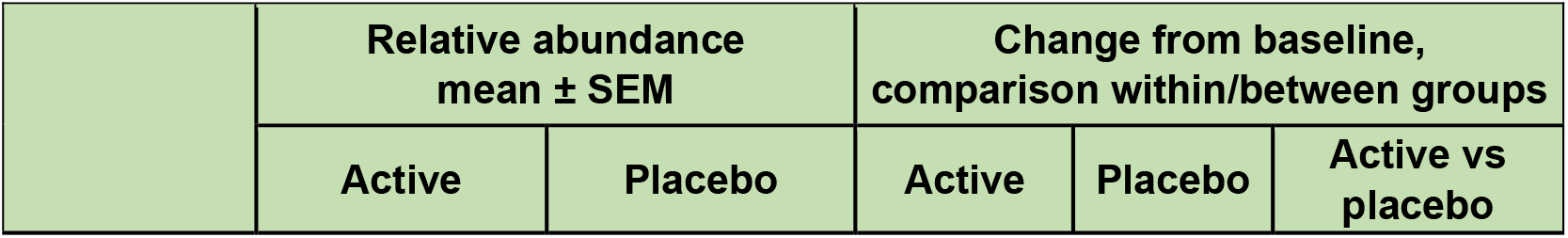
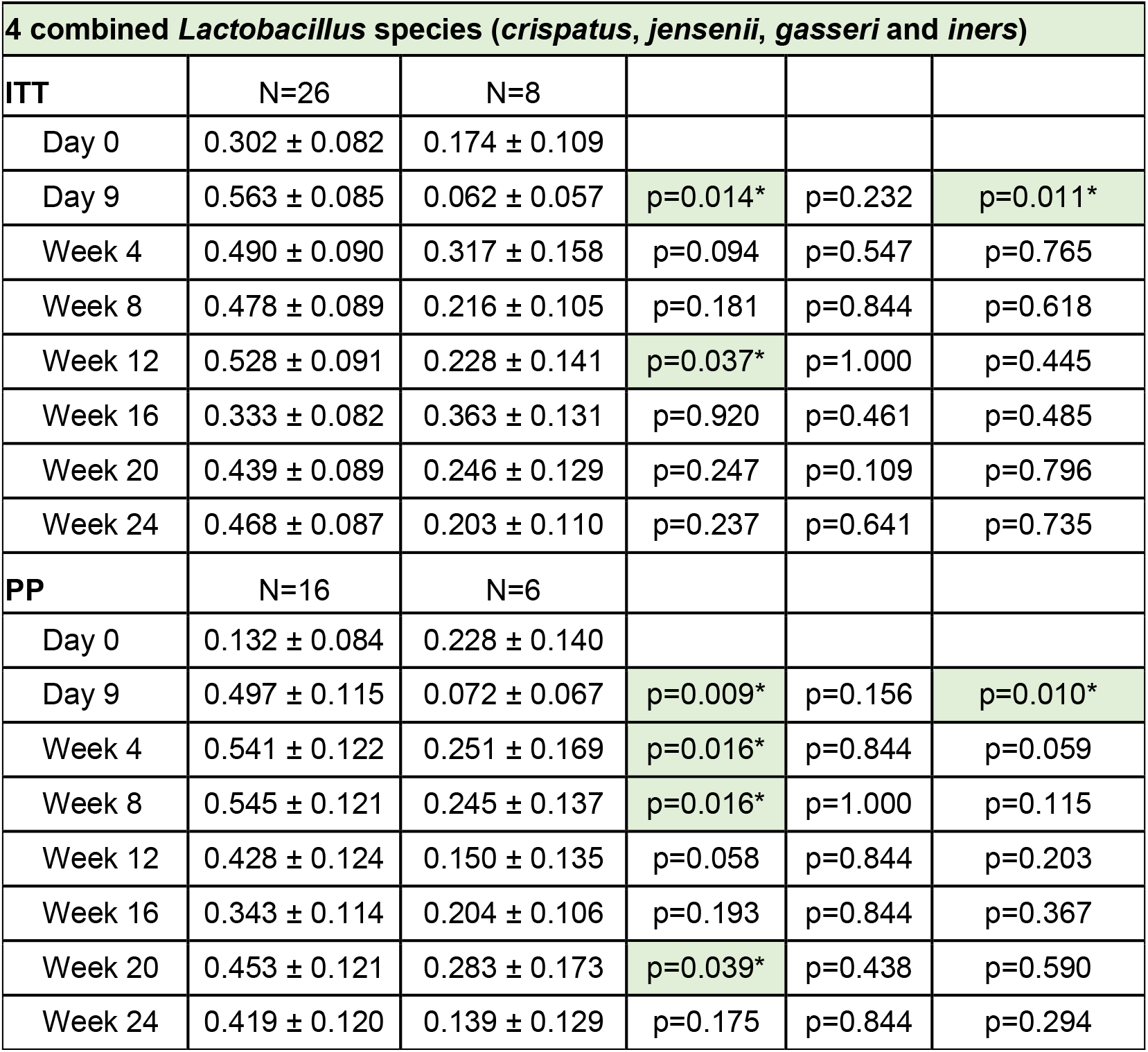
Combined relative abundance of vaginal lactobacilli (*crispatus, iners, jensenii, gasseri*) at each time point from Day 0 to Week 24 after the VMT intervention in the ITT and PP groups. Data are shown as mean ± SEM. P values were calculated using the Wilcoxon signed-rank test and are shown for the Active group compared to Day 0, the Placebo group compared to Day 0, and the Active group compared to the Placebo group at that timepoint. Asterisks indicate values p < 0.05.

| | Relative abundance<br>mean ± SEM | | Change from baseline,<br>comparison within/between groups | | |
| --- | --- | --- | --- | --- | --- |
|  | Active | Placebo | Active | Placebo | Active vs placebo |

**Table 2.** Combined relative abundance of vaginal lactobacilli (*crispatus*, *iners*, *jensenii*, *gasseri*) at each time point from Day 0 to Week 24 after the VMT intervention in the ITT and PP groups.
| 4 combined *Lactobacillus* species (*crispatus*, *jensenii*, *gasseri* and *iners*) |  |  |  |  |  |
| --- | --- | --- | --- | --- | --- |
| ITT | N=26 | N=8 |  |  |  |
| Day 0 | 0.302 ± 0.082 | 0.174 ± 0.109 |  |  |  |
| Day 9 | 0.563 ± 0.085 | 0.062 ± 0.057 | p=0.014* | p=0.232 | p=0.011* |
| Week 4 | 0.490 ± 0.090 | 0.317 ± 0.158 | p=0.094 | p=0.547 | p=0.765 |
| Week 8 | 0.478 ± 0.089 | 0.216 ± 0.105 | p=0.181 | p=0.844 | p=0.618 |
| Week 12 | 0.528 ± 0.091 | 0.228 ± 0.141 | p=0.037* | p=1.000 | p=0.445 |
| Week 16 | 0.333 ± 0.082 | 0.363 ± 0.131 | p=0.920 | p=0.461 | p=0.485 |
| Week 20 | 0.439 ± 0.089 | 0.246 ± 0.129 | p=0.247 | p=0.109 | p=0.796 |
| Week 24 | 0.468 ± 0.087 | 0.203 ± 0.110 | p=0.237 | p=0.641 | p=0.735 |
| PP | N=16 | N=6 |  |  |  |
| Day 0 | 0.132 ± 0.084 | 0.228 ± 0.140 |  |  |  |
| Day 9 | 0.497 ± 0.115 | 0.072 ± 0.067 | p=0.009* | p=0.156 | p=0.010* |
| Week 4 | 0.541 ± 0.122 | 0.251 ± 0.169 | p=0.016* | p=0.844 | p=0.059 |
| Week 8 | 0.545 ± 0.121 | 0.245 ± 0.137 | p=0.016* | p=1.000 | p=0.115 |
| Week 12 | 0.428 ± 0.124 | 0.150 ± 0.135 | p=0.058 | p=0.844 | p=0.203 |
| Week 16 | 0.343 ± 0.114 | 0.204 ± 0.106 | p=0.193 | p=0.844 | p=0.367 |
| Week 20 | 0.453 ± 0.121 | 0.283 ± 0.173 | p=0.039* | p=0.438 | p=0.590 |
| Week 24 | 0.419 ± 0.120 | 0.139 ± 0.129 | p=0.175 | p=0.844 | p=0.294 |

**Figure 3.**
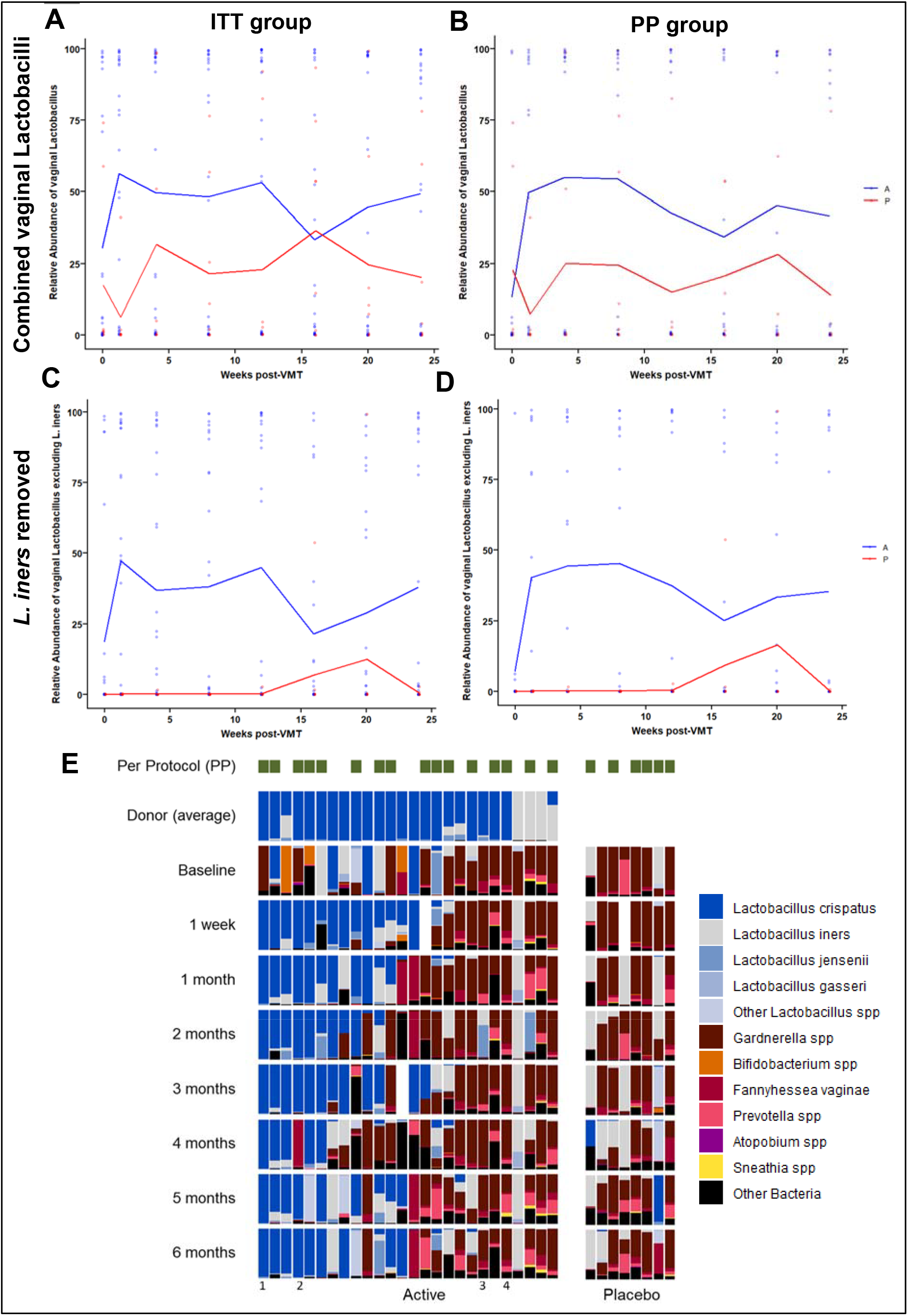
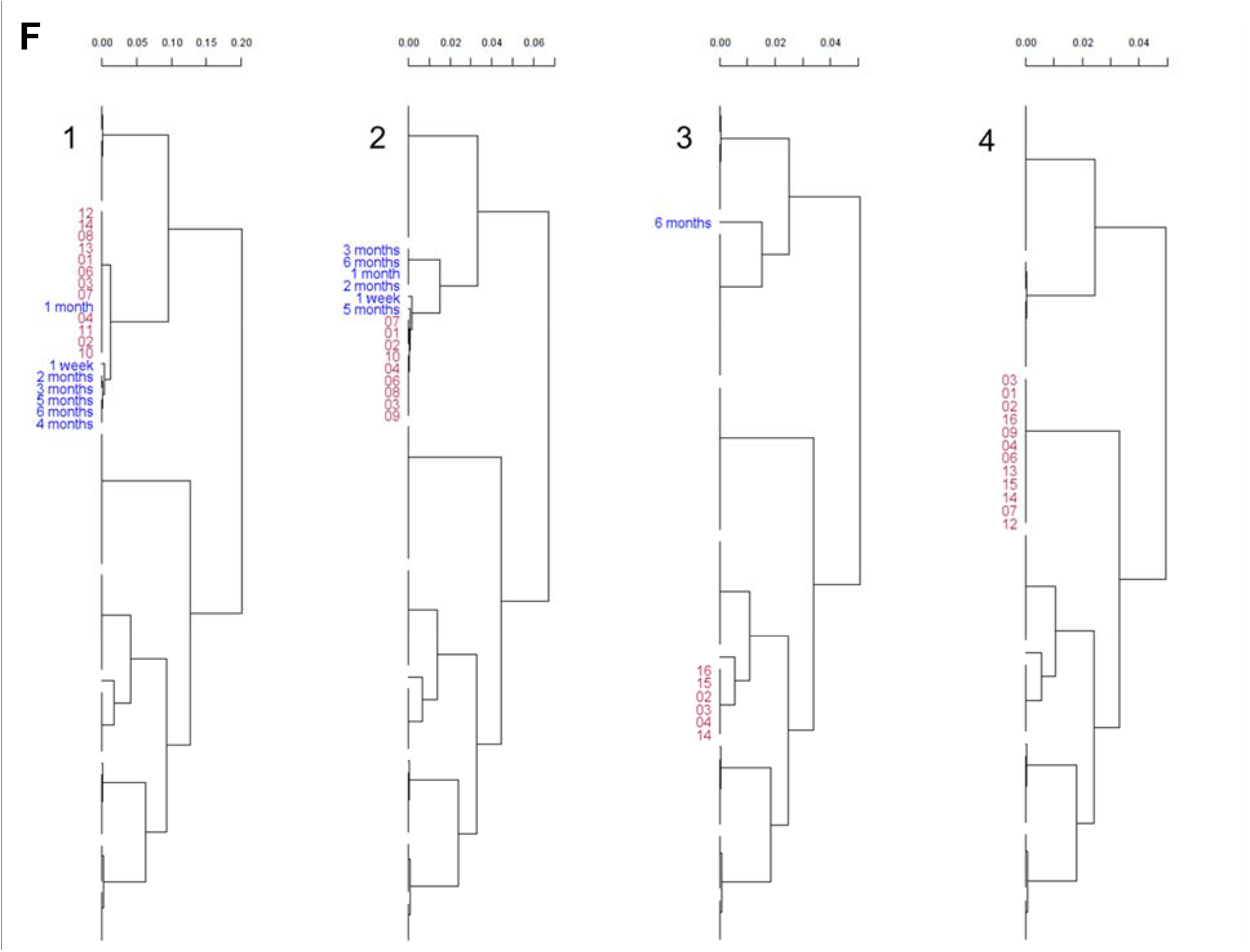
Combined relative abundance of vaginal *Lactobacillus* (*crispatus, iners, jensenii, gasseri*) over time after the VMT intervention in the (A) ITT group (Active n = 26, Placebo n = 8) and the (B) PP group (Active n=16, Placebo n = 6). Separately, participants in the (C) ITT group excluding dominance by *L. iners* in either the donor or recipient (Active n = 22, Placebo n = 8) and the (D) PP group excluding dominance by *L. iners* in either the donor or recipient (Active n = 8, Placebo n = 6). Individual data points represent individuals in the Active (blue) and Placebo (red) groups, and the colored lines represent the mean. (E) Relative abundance of vaginal microbiota observed in each Recipient in the Active and Placebo arms over time. The average relative abundance of bacteria in the 3 Donor CVS samples used for VMT is shown for each recipient in the Active arm. The green blocks at the top of the plot indicate which participants were included in the PP analysis. The numbering 1-4 at the bottom indicates the recipients selected for the engraftment analysis. (F) Dendrograms constructed using hierarchal clustering of Manhattan dissimilarity matrices on *L. crispatus* SNV profiles of 4 sets (labels 1-4 correspond to the labels in (E)) of longitudinal recipient samples compared against all donor samples used in the study. Blue text designates samples containing sufficient *L. crispatus* for SNV analysis at the specified time post-VMT. Red text designates donor samples obtained from the donor that was used for that recipient’s VMT.

### Host biomarker analyses

To determine whether the restoration of *Lactobacillus*-dominance also caused quantitative changes in host biomarkers, transcriptomic analysis was performed on CVS before and after VMT. In **Figure 4A**, the relative change in expression of genes involved in the complement pathway is shown at 8 weeks after VMT and grouped according to the percentage change in the relative abundance of combined vaginal *Lactobacillus* species. Principal coordinate analysis showed a general grouping of CVS samples with increased *Lactobacillus* compared to baseline (**Figure 4B**), and when plotting the PC1 value against the change in *Lactobacillus*, there was a clear trend for both samples with increased and decreased relative abundance (**Figure 4C**). Mapping of the genes involved in the complement pathway showed that many classical genes, such as C1, C2, C3, and C5 were downregulated when *Lactobacillus* dominance was restored, while the inhibitor of the complement pathway, CD59, was upregulated (**Figure 4D**). When assessing a panel of proteins in the plasma of study participants, there were no significant changes in any direction noted for any group (not shown).

**Figure 4.**
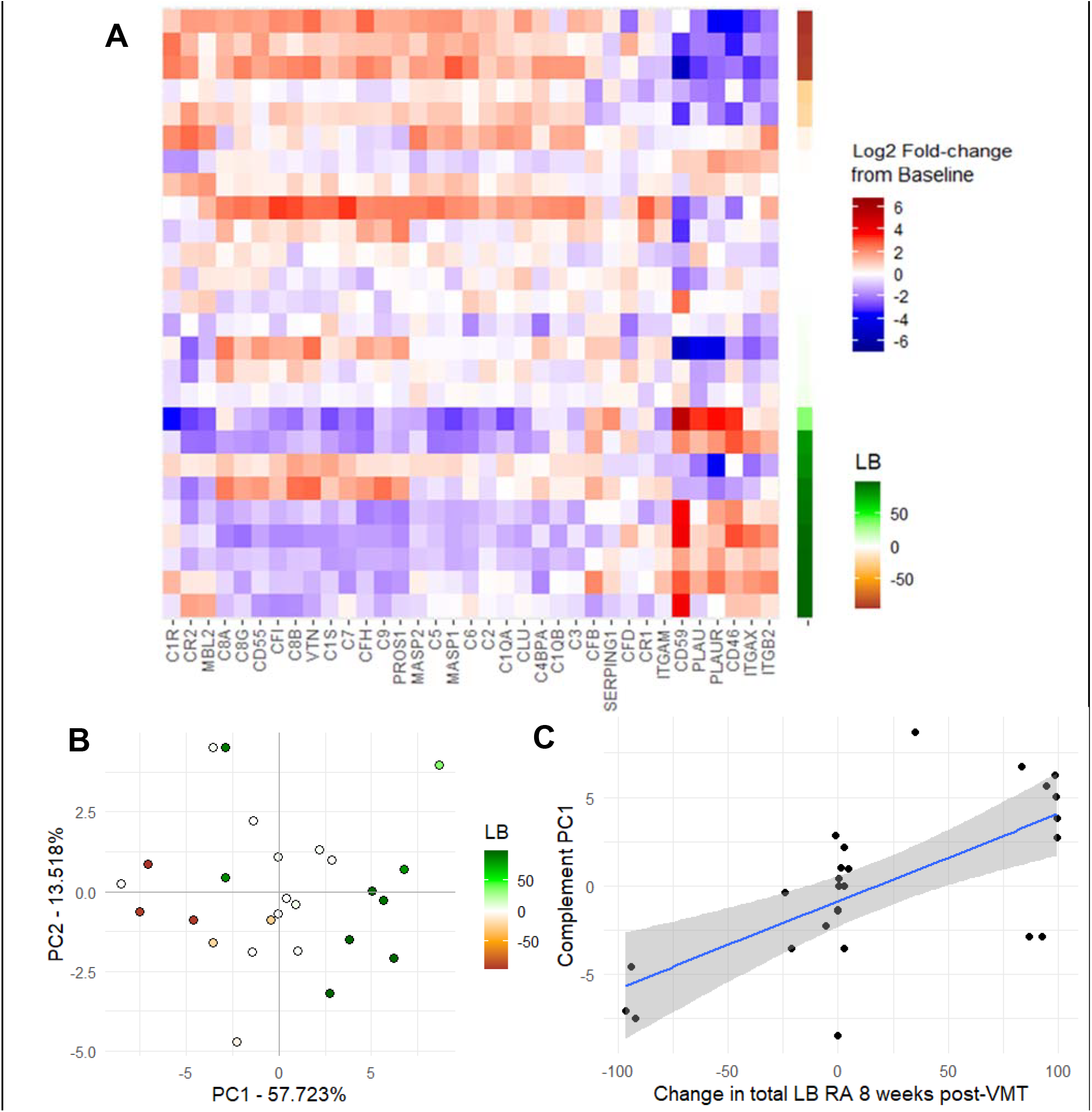
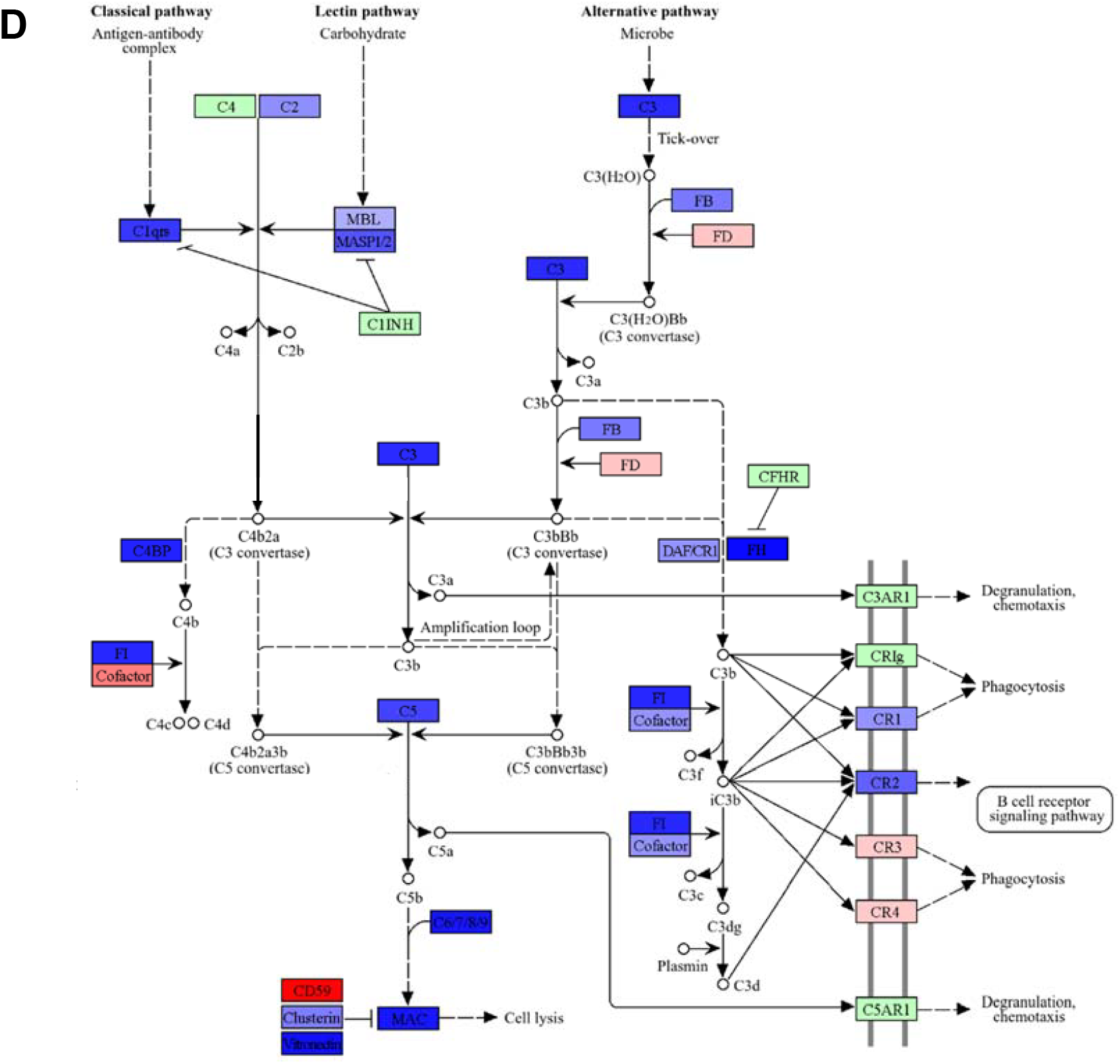
(A) Relative gene expression shown as log2 of the fold change at 8 weeks after VMT compared to baseline. Increased expression is shown in shades of red, and decreased expression is shown in shades of blue. Each gene in the complement pathway is shown labeled at the bottom. Each row represents an individual recipient. The recipients are arranged based on the relative change in relative abundance of total vaginal lactobacillus (LB) species, where shades of green indicates an increase, and shades of burnt orange indicate a decrease. (B) Principal coordinate analysis of the complement gene expression changes in each recipient at 8 weeks after VMT compared to baseline. Each point represents an individual recipient, and the color gradation from green to burnt orange shows the relative change in combined vaginal lactobacillus (LB) species at 8 weeks post VMT compared to baseline. (C) Plot comparing the PC1 coordinate value for each recipient compared to the relative change in total LB at 8 weeks post VMT compared to baseline. A best fit line with the standard error shaded is shown. The points were significantly correlated, p = 0.0003.(D) Cellular mechanism map including the complement-related genes in the analysis for recipients that had an increase in total vaginal lactobacilli at 8 weeks after VMT compared to baseline. Each box represents a related gene. Shade of blue indicate a decrease in expression, while shades of red indicated an increase in expression. Expression of the genes marked in green was essentially unchanged.

## Discussion

Vaginal dysbiosis has been associated with increased prevalence of acute and chronic inflammatory conditions affecting the uterus, ovaries, and bladder, including pelvic inflammatory disease (PID)/endometritis, idiopathic and secondary infertility, endometriosis, polycystic ovary syndrome, preterm birth, and urinary tract infections^36,37^. Here, together with the accompanying paper by Wrønding et al., we provide the first evidence for a causal effect of engraftment of *Lactobacillus*-dominated vaginal microbiota from donor CVS leading to a shift in expression of elements in the inflammatory pathway that is consistent with a reduction in the inflammatory state. Replacement of dysbiosis-associated bacteria with a *Lactobacillus*-dominated vaginal community and resetting the immunological tone of the reproductive tract has the potential to impact a wide range of reproductive and women’s diseases and conditions.

In the VMT study by LevSagie et al., five patients suffering from four or more symptomatic episodes of BV during the previous year, despite repeated and prolonged antibiotic treatment, were treated with antibiotics followed by freshly collected, *Lactobacillus*-dominated vaginal secretions^34^. While promising, there was no placebo arm to characterize the effect of the antibiotic treatment alone. Furthermore, the study did not analyze the microbiome at the strain-level to confirm engraftment with donor bacteria, and the donor samples were collected fresh and used immediately. In a case study of a patient with BV symptoms and recurrent pregnancy losses, we used frozen and banked donor material from the donor program described here and were able to confirm donor strain engraftment using SNV analysis^35^. From a safety and scalability perspective, the ability to freeze and store CVS samples to allow for confirmatory donor testing and without a loss in efficacy is important. The field of FMT evolved in a similar direction, starting from timing the collection, processing, and dosing of donor material to occur on the same day, and then moving to the storage and large-scale screening and banking of donor material^38^. The methodology described here is based on the methods proposed by DeLong et al.^39^ and includes full confirmatory donor testing and the implementation of rigorous quality control and product release criteria that facilitate scale-up and widespread production.

In the companion paper by Wrønding et al., VMT was less successful in a recipient population than what we describe here. The reduced efficacy could be due to the lower dosage (single doses of VMT were given to the recipients, in contrast to the three consecutive doses here), and the inclusion of symptomatic individuals. However, Wrønding et al. identified a procedural improvement by adding an antiseptic pretreatment prior to VMT, which led to successful treatment in 5 out of 10 women in which the intervention previously failed. Taken together, both studies demonstrate the potential of VMT-based interventions.

The development of antimicrobial resistance (AMR) in BV-associated bacteria is a growing concern, particularly with the repeated use of antibiotics in refractory cases and prolonged use of maintenance prophylactic regimens^40^. The high rates of recurrence of BV symptoms after antibiotic treatment combined with the concern of AMR places clinicians in a difficult position without alternative treatment options. Other experimental chemical treatments, such as boric acid and lactic acid, may be employed, but have not been shown to have long-term benefits^41^. LACTIN-V is a pioneering vaginal live-biotherapeutic product containing the CTV-05 strain of *Lactobacillus crispatus*^42^ and has been shown to provide a delay in the recurrence of symptomatic BV, though the effect wanes once dosing stops, likely because these either include non-vaginal bacteria or insufficiently capture the key bacterial strains to mimic VMT engraftment potential^32,43^. Finally, the stable engraftment of donor *Lactobacillus* species demonstrated here with VMT was achieved without antibiotics. Such an approach may help circumvent issues related to repeated use of antibiotics and development of AMR. Moreover, an antibiotic-free approach may be particularly desirable for correction of asymptomatic dysbiosis in conditions related to fertility and pregnancy.

Recent evidence is mounting that even asymptomatic dysbiosis detected using molecular or staining techniques is associated with increased local inflammation^5,28^. Indeed, we demonstrated using a highly sensitive, comprehensive gene expression characterization method that there was differential expression of genes involved in the complement pathway in CVS from women with *Lactobacillus*-dominated microbiota compared to women with asymptomatic dysbiosis. Importantly, the gene expression profiles in the CVS of recipients with an increase in vaginal lactobacilli at 8 weeks after VMT globally led to up- or downregulation of the complement associated genes that was consistent with *Lactobacillus*-dominance. Our human intervention study provides strong evidence that the vaginal microbiota drives immune activation as here demonstrated through regulation of expression of genes in the complement pathway in innate immunity. These findings were corroborated by Wrønding et al. To our knowledge, these studies are the first evidence of the causal relationship between the vaginal microbiota and the local inflammatory state, which goes beyond previously shown associations. Activation of the complement pathway has already been implicated in women at risk of spontaneous preterm birth^29^. The potential positive implications of successful VMT in women with inflammatory conditions that are associated with vaginal dysbiosis, including various fertility and obstetric issues, should be explored.

Our study is not without limitations. Due to the geographical location of the clinical trial, the recipient participants were mainly Caucasian, which limits generalizability to black and Hispanic women, who are more likely to suffer from BV^44,45^. As women with dysbiosis often cycle between states where the vaginal microbiota is dominated by *L. iners*, spontaneous conversion to *L. iners* in the Placebo group impacted the primary outcome measure of combined vaginal *Lactobacillus* after the intervention. However, *L. iners* was noted to have the broadest strain variability, and thus the role of *L. iners* in reproductive tract health remains to be elucidated^46^.

Further, due to COVID-related logistical issues and a high number of the women being screened for participation as a recipient having *Lactobacillus*-dominated microbiota, the sample size was under the targeted number. In addition, our study was insufficiently powered to identify microbiome features in donors and/or recipients that predict engraftment success. We did not observe a “super donor” in our cohort, suggesting that features in both donor and recipient may be important for engraftment success. Overall, the studies described herein describe key fundamental learnings toward the potential of VMT, as well as key methodological foundations to support future larger scale intervention studies in a range of conditions and diseases affecting the female reproductive tract.

## Methods

For detailed methods on donor participant recruitment, sample collection, processing, and analysis, and characterization methods applied to both donor and recipient samples, see the Supplementary Information. The timeline and procedures followed by potential donors are represented graphically in **Figure 1A**.

### Study information

Donor CVS material was collected, processed, and stored by Freya Biosciences in Copenhagen, Denmark in the time period from 2021-2022. The VMT intervention study was conducted as a single-site, randomized, double-blind, placebo-controlled, parallel-group study by the contract research organization Atlantia Clinical Trials (Cork, Ireland) in the time period from November 2021 to October 2022. The study was conducted in accordance with the ethical principles in the Declaration of Helsinki and Good Clinical Practice. The study protocol and related documents describing all study procedures and the donor program were all approved by the Clinical Research Ethics Committee of the Cork Teaching Hospitals (Cork, Ireland) before the study was initiated. The study was registered at ClinicalTrials.gov prior to enrolling participants (NCT05114031).

### Study objectives

The primary objective of the intervention study was to evaluate the change in the vaginal microbiome composition in women with asymptomatic vaginal dysbiosis over the approximately 6-month follow-up period after receiving the VMT intervention. A secondary objective was to characterize the presence of *Lactobacillus* strains originating from the donor CVS samples in the recipients in the active arm after VMT. The safety profile of the VMT intervention was assessed by recording adverse events, vital signs, clinical chemistry and hematology in blood samples, and monitoring of key infections in blood and vaginal swab samples.

### Recipient participants

The participants were all carefully informed about the study and the related procedures before they signed the informed consent form and were screened for participation. The main inclusion criteria for recipients undergoing VMT were: Healthy, pre-menopausal women with age between 18 and 45 years, free of vaginal symptoms that had a combined <10% total relative abundance of *Lactobacillus* spp. and a combined >20% relative abundance of *Gardnerella* + *Atopobium + Prevotella* based on metagenomic sequencing of a vaginal sample obtained at the screening visit. The main exclusion criteria were pregnancy, breastfeeding, current infections including HIV, *Candida* species, BV, *Chlamydia, Gonorrhoea, Mycoplasma genitalium, Trichomonas vaginalis* and/or recent use of systemic of vaginally applied antibiotics.

### Intervention sample size

In one of the few studies investigating the change in vaginal microbiome composition via VMT, it was found that 83% of women in the active arm had a *Lactobacillus*-dominated vaginal microbiome at follow-up^34^. Based on this study, a study describing spontaneous conversion of asymptomatic dysbiosis to *Lactobacillus*-dominance^47^, and a 3:1 ratio of VMT:placebo, a sample size of 40 subjects was calculated to demonstrate significant efficacy at 90% power.

### Intervention study design

The recipient study visits were structured as described in **Figure 1A**. The CONSORT diagram summarizing the recipient study is shown in **Figure 1B**. After the screening visit where all criteria for participation were checked, a baseline visit (Visit 2), was scheduled to be performed around day 8 of the following menstrual cycle. Besides all baseline assessments, the participants were randomized to active VMT or placebo intervention in a 3:1 ratio, and the first VMT was performed. Two additional doses of either VMT or placebo were performed on the subsequent two days for a total of three doses. In 18 of the participants, dosing was initiated later than day 8 of the menstrual cycle, but the three doses were always given on subsequent days. The first follow-up visit (Visit 5) occurred 1 week after the last dose was administered. Visit 6 occurred 4 weeks (1 menstrual cycle length) after the first dosing, and visits 7-11 were scheduled to occur with approximately 4 weeks spacing thereafter **(Figure 1A**).

For detailed methods describing the assessment of the primary and secondary objectives and biomarker characterization, see the Supplementary Information.

## Data Availability

The datasets generated during and/or analyzed during the current study are available from the corresponding author on reasonable request.

## Acknowledgements

We acknowledge Solrun Breddam, Marianne Wettlaufer, and Tina Devitt for technical support.

## Contributions

EFB, BM, RR, TGR, CA, UKB, PC, ABT, FZ, TD, FPM, JETHV, LME, HSN, TW contributed to the study conception, design, and supervision. EFB, BM, KD, MAR, HBJ, AMA, MJS, RLM, HCI, HSN, TW, PC, MO, SP, and SD were responsible for data acquisition, analysis, and interpretation JAN, TD, BM, FM were responsible for cohort recruitment and participant intervention. EFB, BM, KD, LME, and JETHV drafted and revised the manuscript. All authors contributed and critically reviewed the final version of the manuscript.

