## Supplementary information for "Antibiotic-free vaginal microbiota transplantation (VMT) changes vaginal microbiota and immune profile in women with asymptomatic dysbiosis – reporting of a randomized, placebo-controlled trial"

Supplementary Methods

Donor recruitment and prescreening

Healthy pre-menopausal women without a history of BV or other infections or conditions affecting the female reproductive tract were recruited. Due to the generally high prevalence of human papilloma virus (HPV) in this age group, study participants were first screened for HPV and vaginal microbiome composition before further in-depth screening. A total of 96 women consented and provided samples for prescreening. For the first 36 women screened, one vaginal swab each was self-collected for HPV testing and microbiome analysis. For the last 60 women screened, a CVS sample was obtained for microbiome characterization instead of a swab. Microbiome analysis from CVS has been shown to be indistinguishable from vaginal swabs, while also providing larger amounts of material for DNA extraction and other characterizations[1]. For the CVS self-collection, a pliable menstrual cup (Flex disc, The Flex Company) was inserted into the vaginal cavity prior to placing in a 50 mL conical tube for centrifugation and collection, as previously described[2].

HPV testing

HPV testing was performed on both donor and recipient samples. DNA extraction was performed using the Qiagen QIAmp DNA mini kit. Extracted DNA from each donor sample was used in the SeeGene Anyplex II HPV28 kit and ran semi-quantitatively on the BioRad CFX96 Dx qPCR machine, which was set up and calibrated for this assay (Triolab, DK). The semi-quantitative method detects 28 different genotypes of HPV, including all known high-risk genotypes. Only women that tested negative for all HPV strains continued the screening.

Microbiome analysis

Microbiome analysis was applied to both donor and recipient samples. Swabs for vaginal microbiome analysis were kept at 4°C for up to 48 hours prior to DNA extraction. CVS samples were diluted with saline to reduce the viscosity, aliquoted, and stored at -80°C prior to DNA extraction. DNA was extracted from both sample types using the Molysis Complete5 kit (MolZym), which uses a differential lysis method to extract microbial DNA and remove human DNA. Shotgun sequencing and bioinformatic analyses were performed by Seqbiome (Cork, IE). All samples were prepared for shotgun metagenomic sequencing according to Illumina Nextera XT library preparation kit guidelines, with the use of unique dual indexes for multiplexing with the Nextera XT index kit (Illumina). Samples were sequenced on an Illumina NextSeq 500 sequencing platform with a v2.5 kit (a 300-cycle kit/150bp PE sequencing), at the Teagasc DNA Sequencing Facility, using standard Illumina sequencing protocols. The quality of the raw sequencing data was assessed using FastQC. Samples that did not meet minimum quality check cutoff or a sequencing depth of at least 4M reads underwent re-sequencing. Shotgun metagenomic sequencing data were then processed through analysis workflow that utilizes Kneaddata wrapper tool. Quality filtering and host genome decontamination (human) were performed utilizing Trimmomatic and Bowtie2. Taxonomic classification of quality filtered reads was further performed using Kraken2 species classifier using a customized version of Genome Taxonomy Database (GTDB) that also includes reference sequences belonging to archaea, fungi, and viral genomes. Kraken2 classifications were then passed to Bracken tool to estimate species level abundance. The vaginal microbiome composition was then defined as relative abundance of bacterial species. *Atopobium* and *Prevotella* species were grouped together and labeled at the genus level. Only women that had >80% relative abundance of *Lactobacillus* species (*L. crispatus, L. iners, L. jensenii, L. gasseri)* and <5% combined relative abundance of selected dysbiosis-associated bacteria (*Atopobium spp., Prevotella spp., B. vaginale, and F. vaginae*) and negative HPV test continued the Donor screening process.

Additional donor testing

To minimize the risk of pathogen transmission to the recipients, potential CVS donors were further screened by a midwife at TFP Stork Fertility for the presence of sexually transmitted diseases and underwent a medical and gynecological examination to assess the presence of other diseases such as cancer or endometriosis. A general health check including medical history and medication usage, demographics, heart rate and blood pressure measurements was performed. Standard clinical diagnostic tests were performed for human immunodeficiency virus (HIV), hepatitis A, B, and C, cytomegalovirus, Treponema pallidum, urinary tract infection, human papilloma virus (HPV), *Chlamydia trachomatis*, *Neisseria gonorroheae*, *Trichomonas vaginalis*, herpes simplex virus 1 and 2, Candida species, *Mycoplasma genitalium*, and *Streptococcus* A, B, C, and G. If all test results were normal/negative, then the participant was able to enter the donor program and provide CVS samples as described below.

Donor CVS sample collection schedule

The timeframe for donor CVS collection was a 40-day period between the two examination and testing visits. Each donor provided 10-15 donations at any time during the 40 days except during menstruation and one day thereafter. The donations were spaced at least 16 hours apart, as CVS samples collected on the same day (two samples 6 h apart or three samples 4 h apart) resulted in increased pH, decreased sample weight, and decreased CFU (not shown). During the donation period, donors needed to adhere to the following restrictions: abstinence from vaginal and anal sexual intercourse; no swimming in lakes, hot-tubs, or swimming pools; no use of intravaginal products (e.g., tampons, soap). At each visit, the donor filled out a questionnaire to affirm adherence to these restrictions together with general health questions. Upon completion of the sampling, the CVS samples were held in quarantine until the donor was subjected to and similarly passed the donor testing described above. Donors were allowed to complete multiple donation periods as long as all inclusion criteria and pathogen testing were completed before and after the CVS donation period. Donors 2, 5, and 7 had a 1 month gap between two rounds of CVS donations, and donor 1 had a 1.5 month gap between two rounds of CVS donations. Donor 6 had 2 months between the rounds and changed from using contraceptive pills in the first round to no hormonal contraceptives in the second round.

Donor CVS collection procedure

Donors self-collected CVS samples in a dedicated, hygienically designed donor room. The donor room was cleaned with 70% ethanol after each donor and subjected to biweekly environmental monitoring to check for Enterobacteriaceae, total aerobic microbial count, yeast, and mold. Each batch of soft, flexible menstrual cups were confirmed to be free of contamination by Enterobacteriaceae. The donor self-collected CVS by pinching and collapsing the menstrual cup and inserting it into the vagina. The cup was left in a longitudinal position for about 10 seconds prior to twisting it along its longitudinal axis while removing it. The donor then placed the cup into a sterile, labeled, pre-weighed 50 mL conical tube. After 15-20 min, this process was repeated a second time with a second menstrual cup, which was placed in a separate sterile 50 mL tube. The CVS was collected by centrifuging for 5 min at 190 x g and ambient temperature. All further steps were conducted in a sterilized biosafety cabinet at ambient temperature. The self-sampling devices were discarded from the tubes and the CVS sample weight was determined. Samples with a total (two tubes combined) weight of less than 200 mg were discarded. Samples in which blood was visibly present were discarded.

Donor CVS processing and storage

The CVS sample in each tube was mixed with 1 mL saline to reduce viscosity, after which the two samples were combined and aliquoted for storage and further testing. The two combined samples were then distributed over several aliquots for quality control and storage. Pilot tests confirmed that aliquotting and storing smaller CVS volumes for stability and other characterizations did not negatively impact quantification of viable bacterial cell counts (not shown). The cryovials containing samples for storage were placed in a CoolCell (Corning) and then placed in a -80°C freezer. After a minimum of 2 hours, the samples were transferred into a regular -80°C storage box labelled 'quarantined'. Samples were released only after all release criteria had been met (see below). The lot of released CVS samples was shipped on dry ice with a temperature logger by overnight courier to the trial site, and the samples were stored at –80° C until thawing for administration.

CVS quality control systems

The sample volumes used for analyses and quality control were kept as low as possible to minimize CVS sample loss. A sample aliquot was used to measure pH with a microelectrode. Technical triplicate measurements were performed, and repeated if the difference between measurements was > 0.2 pH units. The sample used for pH measurement was discarded after measurement. The retention vial was maintained for at least 1 year after administration for safety checks as required. DNA was extracted from an aliquot of every sample for microbiome analysis (described above). DNA from the first and last CVS sample from each donor was subjected to a qPCR analysis to check for the absence of multi-drug resistance genes using the SeeGene Allplex Entero-DR qPCR assay kit on a BioRad CFX96 Dx qPCR machine calibrated for the assay. This kit allowed for single or multiple detection of carbapenemase genes (NDM, KPC, OXA-48, VIM, IMP), extended spectrum beta-lactamase (ESBL) genes (CTX-M), and vancomycin resistance genes (VanA, VanB). Microscopy was performed on every CVS sample to look for evidence of clue cells, white blood cells, sperm cells, and yeast hyphae. The absence of sperm cells was confirmed using acid phosphate paper. Viability of samples dominated by *L. crispatus*, *L. jensenii,* or *L. gasseri* was quantified by colony-forming units (CFU) count on de Man, Rogosa and Sharpe (MRS) agar plates or with viable cell count (VCC). For CVS dominated by *L. iners*, viability was tested using BacLight live/dead staining (ThermoFisher Scientific) and counting the viable cells/mL in a Thoma cell counting chamber under the microscope. Within one week after a CVS sample was dosed to a recipient, a stability vial was used to test bacterial cell viability at the time of administration.

Donor CVS sample preparation and administration

Frozen CVS samples were first thawed by transferring from -80°C to room temperature for 40 minutes. Thereafter, the sample was placed in an incubator at 37°C for 30 minutes. The tube containing the sample was then gently inverted 5 times for mixing. The sample was drawn up into a 5 mL syringe and applied intravaginally to the recipient lying in the lithotomy position using an insemination catheter attached to the 5 mL syringe. The recipient was instructed to remain lying down in a horizontal position for 30 minutes following the sample being inserted intravaginally.

Recipient intervention data collection

Participants had vaginal swabs analyzed for *Chlamydia trachomatis*, *Neisseria gonorrhoeae*, *Trichomonas vaginalis*, *Mycoplasma genitalium*, herpes simplex virus 1 and 2, *Candida* spp., and HPV at screening, baseline, and 4 weeks after VMT. Blood samples were collected at the same visits for analysis of HIV 1 and 2, hepatitis A, B, and C, cytomegalovirus, and *Treponema pallidum*. Hepatitis A and cytomegalovirus immunoglobulin G (IgG) antibodies, indicating previous infection and immunity, were detected at baseline and week 4 in both groups. Urine samples were collected for assessment of urinary tract infections and pregnancy testing. A broad panel of clinical chemistry or hematology parameters were measured in venous blood samples collected at screening, baseline and 1 menstrual cycle length into the follow-up period. Vitals signs (blood pressure, heart rate, temperature) were measured at screening, baseline and all follow-up visits. Adverse events (AEs) were recorded from the baseline visit and throughout the follow-up period.  Some AEs were defined in the Safety Monitoring Plan as Adverse Events of Special Interest (AESIs). The following events were defined as AESIs: any gynecological conditions, specifically sexually transmitted infections and other infections, reported from day 0 to week 8 (Visit 7), as well as any abnormal vaginal bleeding, including changes in menstruation, irrespective of timing. Detection of *Ureaplasma* by qPCR was included in the panel of tests and in the definition of AESI, though there are no established clinical criteria or diagnostic paradigms associated with the detection of *Ureaplasma*. The presence of *Ureaplasma* was already common at baseline, where 56% (19 of 34 subjects) of the trial population had vaginal samples with a presence of *Ureaplasma*. Any gynecological condition that occurred from Week 8 to Week 24 (Visit 7 to 11) into follow-up was recorded as an AE. “Related” is a subjective assessment by the sub-investigator, so if a participant presents with a symptom and complains that she never experienced this before the study, the sub-investigator may enter it as a (possible/probable) related AE. While if the participant said that it is a common returning symptom for her, it would likely be listed as unlikely related.

Recipient intervention randomization and masking

Before the study was conducted, the allocation of recipient participants in a 3:1 active:placebo ratio was planned according to a randomization list. Randomization blocks of n = 4 were used throughout, and the study site and everyone involved in data generation and/or analyses of every obtained sample were kept blinded to the randomization. At screening, potential recipient participants were assigned a 3-digit screening number according to their chronological entry into the study. If an individual was found eligible and enrolled for study participation, they were allocated a randomization number in a sequential manner by study site staff after all baseline assessments performed at visit 2 had been performed.

The study product was contained in identical cryovials with identical labeling, except for a masked 5-6 digit sample number. All study participants, the clinical team, and everyone involved in the conduct of the study and generating data were blinded during the entire study until database lock and signature of the request for the unblinding document. An emergency unblinding procedure with emergency code-break envelopes was established to allow the investigator the option of disclosing the product assignment for any individual participant if clinical circumstances required such unblinding, though this was not required during the study.

Primary objective

The primary objective of evaluating the change in the vaginal microbiome composition over the approximately 6-month follow-up period in the recipients was conducted for both the ITT and PP study populations. Sequencing was performed on CVS samples as described above. The combined relative abundance of vaginal *Lactobacillus* species (*L. crispatus, L. iners, L. jensenii, L. gasseri)* was calculated for each recipient at each follow-up visit. Due to the ambiguous role of *L. iners* in vaginal health, the characterization was also done with exclusion of *L. iners* and the recipients that received *L. iners*-dominated donor CVS.

*L. crispatus* SNV profile comparison

After quality filtering and host genome decontamination *L. crispatus* reads in each sample were aligned to a reference genome (GCF_02901145.1) using Burrows-Wheeler Aligner (0.7.17) and SNV profiles were generated using metaSNV (2.0.4) requiring an average genome coverage depth of 3, and default settings. A Manhattan dissimilarity matrix was calculated for each set of recipient samples along with all donor samples used in the study. Dendrograms were generated for each recipient set using hierarchical clustering in R.

CVS processing for biomarker analysis

CVS samples used for biomarker analysis were immediately placed on ice when received and kept cold throughout the entire processing procedure. Cold instead of room temperature sterile saline was used, and samples were frozen to -80°C without the use of a CoolCell. Instead of adding 1 ml sterile saline per sample, 2.5 µL was added per mg sample so all samples were normalized for weight prior to analysis. The quality checks were omitted (CFU, pH, sperm cell check), except microbiome sequencing.

RNA extraction from CVS

Extraction of total RNA was performed from 100 µL of CVS using the NucleoSpin® 96 RNA kit (Machery Nagel, cat. no. 740709.4). The extractions were done manually in a centrifuge. In brief, the samples were initially thawed on ice, and then 600 µL of buffer RA1 was added to each sample. The lysate was then homogenized in a Tissuelyser II instrument (Qiagen) using two steel beads and shaking at 20 Hz for 2 x 2 min. 700 µL of buffer RA4 was added to the lysate. After mixing by pipetting the lysate from each sample was transferred to the individual wells of the 96 well NucleoSpin® RNA Binding Plate for binding of the nucleic acids. The remaining part of the extraction was performed following the NucleoSpin® 96 RNA protocol (v. February 2023/Rev. 10). The subsequent RNA was eluted in 75 µL of RNase-free $H_2O$.

RNA quality control, normalization, and concentration

The RNA concentration and integrity (measured as RIN values) of all extracted RNA samples were analyzed on an Agilent 4200 Tapestation system (Agilent Technologies). The concentration was normalized to 20 ng/µL based on DV200 values (percentage of fragments >200 nucleotides). Samples with a concentration below 20 ng/µL were concentrated using speedvaccing and then normalized to 20 ng/µL.

Analysis of RNA samples on Nanostring

The extracted RNA was analysed on the nCounter Human Immunology v2 panel (Nanostring, cat. no. XT-CSO-HIM2-12) including a custom panel consisting of 55 targets following the CodeSet Hybridization Setup manual (MAN-10056-03 (January 2020)). 100 ng of total RNA was used as input for the hybridizations. The hybridized RNA:probe complexes were transferred to the nCounter Prep Station (Nanostring) for post-hybridization processing. Scanning was performed on the nCounter Digital Analyzer (Nanostring) using the FovCount 555 value setting. The generated raw data were then analyzed in the nSolver v. 4.0 software (Nanostring). The data were normalized for input variability by scaling the sample values to the geometric mean of the most stable reference genes, which were determined by the geNorm algorithm[3].

Technical validation of sample pH effect on Nanostring

The validation of the effect of CVS pH on the Nanostring results was performed on 4x100 µL CVS per sample for 5 eubiotic and 5 dysbiotic samples. The analysis was performed on recipient CVS with 2.5 µL saline per mg CVS, i.e., normalized for weight. Sample pH was measured and modified as follows. For each sample, 100 µL CVS was analyzed unmodified, 100 µL sample had the pH flipped (pH lowered to 3.8-4.1 for dysbiotic samples using lactic acid, pH increased to 4.7-5.0 for eubiotic samples using NaOH), and 100 µL sample was neutralized to pH 6.5 (+/- 0.5) using neutralization buffer PFN. Furthermore, some CVS from each sample was diluted 2x and 5x with normal sterile saline (0.9% NaCl) to a total volume of 100 µL each for Nanostring analysis. Data analysis confirmed that changing the sample pH did not affect the overall gene expression profiles for the samples (not shown).

Preparation and proteomic analysis of plasma samples

Plasma samples were thawed and vortexed at 2000 rpm for 30 s and spun down at 400 x *g* for 1 min at room temperature. Relative protein abundance was quantified using Proximity Extension Assay (PEA) from Olink Proteomics AB, Sweden[4]. In brief, 1 µL of sample was incubated with 92 antibody pairs labeled with DNA oligonucleotides. Dual binding to the target proteins in proximity of matching antibody-probes resulted in hybridization of the oligos, and a PCR target sequence was formed. Subsequently the target sequence was detected and quantified using a Fluidigm 96.96 IFC and the Fluidigm BioMark HD platform. The PEA readout was Normalized Protein Expression (NPX), which is an arbitrary unit on log2 scale where a high NPX corresponds to a high protein abundance. External and internal controls were included in the PEA assay to monitor assay performance and adjust for intra- and inter-run variation. Four internal controls were added to each sample to monitor the quality of assay performance in each step, i.e. antibody binding, extension and detection step. Negative controls and inter-plate positive controls were run in triplicate on each plate[4]. Assay specific limit of detection was calculated as 3 times the standard deviation over the background signal. Normalization between plates was performed using intensity normalization.

Statistical methods

Analyses for the primary endpoint were conducted using IBM SPSS Statistics Software Version 28.0 (Clinical objectives) and R Project for Statistical Computing Version 4.1 (Microbiome objectives), using validated and tested analysis scripts, as appropriate. Wilcoxon signed-rank test was used to determine between-product differences in absolute change in relative abundance of combined vaginal lactobacilli (either combined crispatus, iners, gasseri, jensenii or combined crispatus, gasseri, jensenii) from Day-0 (Baseline) to Day-9 (Post-VMT Visit) – Week-24. Statistical tests were conducted with a 2-sided hypothesis with a significance level of 5%.

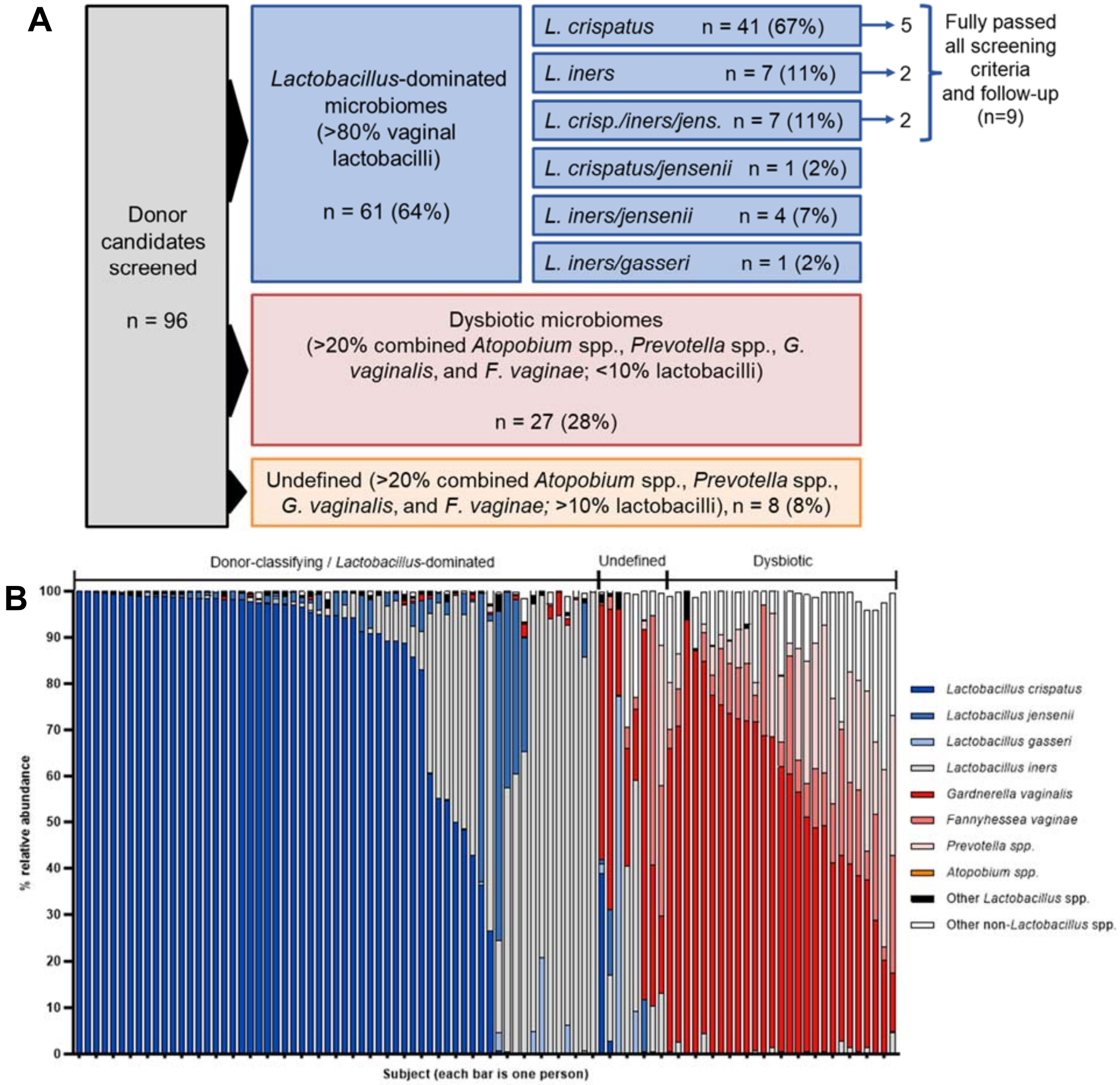


**Supplementary Figure 1**. (A) Schematic representation of the cohort screened for participation as donors. A total of 96 women were screened, 64% (61) of which had *Lactobacillus*-dominated microbiota in their CVS. Of these women, a total of 9 passed the final screening criteria and follow-up testing. (B) Relative abundance of taxa in the vaginal microbiota observed in CVS samples in the donor screening cohort as characterized by sequencing. Each bar represents an individual CVS sample from an individual screening participant. The bars are grouped based on whether they categorized as *Lactobacillus*-dominated, dysbiotic, or undefined.

| **Screening test** | **n** |
|---|---|
| Human immunodeficiency virus (HIV) | 0 |
| Hepatitis A/B/C | 0 |
| Cytomegalovirus (CMV) IgM | 0 |
| Treponema pallidum | 0 |
| Urinary tract infection (UTI) | 2 |
| Human papilloma virus (HPV) | 13 |
| Chlamydia trachomatis | 1 |
| Neisseria gonorrhoeae | 0 |
| Trichomonas vaginalis | 0 |
| Herpes genitalis | 0 |
| Mycoplasma genitalium | 1 |
| Streptococcus A/B/C/G | 4 |
| Candida | 7 |
| Other (e.g., abnormal gynecological findings) | 4 |
| Total passed | 12 |
| Failed during donations or at follow-up | 3 |
| **Total fully passed/approved donors** | **9** |

**Supplementary Table 1**. Summary of key testing failures for potential donors that had *Lactobacillus*-dominated microbiota and tested negative for HPV in the first screening step (n = 38 out of 96). The presence of various pathogens was the most common cause of screening failure. A total of 12 women passed this second screening step, of which 9 also passed follow-up testing and their CVS was used for the VMT intervention.

| | Optimal (n=61) | | Approved donors (n=9) | | Dysbiotic (n=27) | | Undefined (n=8) | |
|---|---|---|---|---|---|---|---|---|
| **Species** | **median** | **IQR** | **median** | **IQR** | **median** | **IQR** | **median** | **IQR** |
| *Lactobacillus crispatus* | 94.75 | 50.05 | 97.42 | 50.16 | 0.00 | 0.00 | 0.03 | 1.08 |
| *Lactobacillus gasseri* | 0.00 | 0.08 | 0.00 | 0.00 | 0.00 | 0.00 | 0.28 | 3.84 |
| *Lactobacillus iners* | 0.84 | 42.20 | 0.84 | 46.48 | 0.34 | 1.11 | 11.47 | 20.79 |
| *Lactobacillus jensenii* | 0.74 | 2.82 | 0.07 | 1.34 | 0.00 | 0.00 | 0.10 | 3.42 |
| **Total vaginal lactobacilli** | **99.26** | **1.44** | **99.62** | **0.37** | **0.34** | **1.10** | **35.76** | **33.35** |
| *Atopobium* spp. | 0.00 | 0.00 | 0.00 | 0.00 | 0.00 | 0.01 | 0.00 | 0.00 |
| *Gardnerella vaginalis* | 0.01 | 0.30 | 0.00 | 0.08 | 61.60 | 31.71 | 27.98 | 39.65 |
| *Fannyhessea vaginae* | 0.00 | 0.00 | 0.00 | 0.00 | 10.70 | 9.75 | 2.63 | 10.11 |
| *Prevotella* spp. | 0.00 | 0.01 | 0.00 | 0.00 | 10.23 | 22.19 | 0.02 | 0.46 |
| **Total selected pathobionts** | **0.03** | **0.50** | **0.01** | **0.08** | **87.21** | **10.10** | **61.83** | **49.59** |
| Other lactobacilli | 0.31 | 0.55 | 0.15 | 0.14 | 0.00 | 0.00 | 0.28 | 0.98 |
| Other species | 0.14 | 0.58 | 0.03 | 0.33 | 12.48 | 10.96 | 5.71 | 13.50 |

**Supplementary Table 2**. Median relative abundances of key bacteria species in CVS samples obtained from women in the donor screening cohort as characterized by sequencing. The columns show the Lactobacillus-dominated group (optimal, >80% Lactobacillus by relative abundance relative abundance), the final group of fully approved donors, the dysbiotic group (>20% combined Atopobium spp., Prevotella spp., G. vaginalis, and F. vaginae; and <10% lactobacilli), and the undefined group (>20% combined Atopobium spp., Prevotella spp., G. vaginalis, and F. vaginae; and >10% lactobacilli). Data shown as means and interquartile ranges (IQR).

| Item | | Min | Max | Average |
|---|---|---|---|---|
| Age | | 22 | 30 | 26.1 |
| BMI | | 20.1 | 35.8 | 27.2 |
| No of donations | | 10 | 15 | 12.8 |
| | | N | | |
| Ethnicity | Caucasian | 7 | | |
| | Asian | 1 | | |
| | Mixed | 1 | | |
| Contraceptive use | none | 3 | | |
| | hIUD | 3 | | |
| | pill | 3 | | |

| | | Ranges (min-max) | | | | Averages | | | | |
|---|---|---|---|---|---|---|---|---|---|---|
| Donor ID | Time window (days) | Weight (mg) | Dose (mL) | pH | Viable cells/dose | Weight (mg) | Dose (mL) | pH | Viable cells/dose (fresh) | Viable cells/dose (at time of VMT) |
| 1 | 28 | 244-1035 | 1.5-1.8 | 3.9-4.4 | 8.6E+06-3.1E+08 | 515 | 1.6 | 4.1 | 1.2E+08 | 1.3E+08 |
| 2 | 36 | 311-918 | 1.4-1.8 | 4.1-4.3 | 1.9E+08-5.4E+08 | 597 | 1.7 | 4.2 | 3.6E+08 | 1.8E+08 |
| 3 | 25 | 628-1778 | 1.7-1.8 | 3.7-3.8 | 5.1E+07-8.4E+10 | 1206 | 1.8 | 3.8 | 1.5E+10 | 2.4E+08 |
| 4 | 31 | 195-486 | 1.3-1.7 | 3.9-4.2 | 7.2E+07-3.4E+08 | 334 | 1.5 | 4.1 | 1.9E+08 | 2.8E+07 |
| 5 | 30 | 340-823 | 1.5-1.8 | 3.7-4.0 | 3.3E+08-1.0E+11 | 525 | 1.7 | 3.9 | 7.4E+09 | 5.2E+08 |
| 6 | 20 | 276-703 | 1.3-1.8 | 3.8-3.9 | 5.0E+07-1.0E+11 | 487 | 1.6 | 3.8 | 1.7E+10 | 2.2E+08 |
| 7 | 16 | 376-710 | 1.6-1.8 | 3.8-4.0 | 3.6E+08-1.0E+11 | 542 | 1.7 | 3.9 | 8.5E+09 | 2.7E+08 |
| 8 | 31 | 532-1593 | 1.6-1.8 | 3.9-4.0 | 4.3E+08-1.2E+09 | 868 | 1.8 | 3.9 | 8.5E+08 | 1.0E+09 |
| 9 | 24 | 233-627 | 1.4-1.8 | 3.9-4.2 | 1.3E+08-6.0E+08 | 446 | 1.6 | 4.0 | 3.0E+08 | 2.0E+08 |
| Average | 27 | | | | | 614 | 1.6 | 4.0 | 5.5E+09 | 3.1E+08 |

**Supplementary Table 3.** Demographics of the donor participants who fully passed all screening and confirmatory testing, and thus, their CVS samples were used in the intervention study. hIUD = hormonal intrauterine device, pill = combined oral contraceptive pills. The number and physical characteristics of the CVS samples obtained from each donor. The range and average values across the samples from each participant are shown in each row, which includes all samples provided by the donor participant (a subset were used for VMT). The bottom row shows averages across all 9 donor participants.

| | Active (n = 26) | | | | Placebo (n = 8) | | | |
|---|---|---|---|---|---|---|---|---|
| | Baseline | | Week 4 | | Baseline | | Week 4 | |
| | n | % | n | % | n | % | n | % |
| **Vaginal swabs** | | | | | | | | |
| Human papilloma virus (HPV) | 14 | 53.8 | 12 | 46.2 | 4 | 50.0 | 4 | 50.0 |
| Candida | 4 | 15.4 | 5 | 19.2 | 1 | 12.5 | 1 | 12.5 |
| Chlamydia trachomatis | - | - | - | - | - | - | - | - |
| Nesseria gonorrhaeae | - | - | - | - | - | - | - | - |
| Mycoplasma genitalium | - | - | - | - | - | - | - | - |
| Trichomonas vaginalis | - | - | - | - | - | - | - | - |
| Herpes simplex virus (HSV) -1 and -2 | - | - | - | - | - | - | - | - |
| **Blood Sample** | | | | | | | | |
| Treponema pallidum | - | - | - | - | - | - | - | - |
| Cytomegalovirus IgG Ab | 12 | 46.2 | 12 | 46.2 | 3 | 37.5 | 3 | 37.5 |
| Cytomegalovirus IgM Ab | - | - | - | - | 1 | 12.5 | 1 | 12.5 |
| Human immunodeficiency virus (HIV) -1 and -2 | - | - | - | - | - | - | - | - |
| Hepatitis A IgG Ab | 11 | 42.3 | 13 | 50.0 | 3 | 37.5 | 3 | 37.5 |
| Hepatitis A IgM Ab | - | - | - | - | - | - | - | - |
| Hepatitis B Core Ab | - | - | - | - | - | - | - | - |
| Hepatitis B S Ag | - | - | - | - | - | - | - | - |
| Hepatitis C Ab | - | - | - | - | - | - | - | - |

**Supplementary Table 4**. Safety analyses focused on testing for infections at baseline compared to week 4 after the intervention in the ITT population. No notable differences were observed in either the Active or Placebo arms.

| | | Related n(%)E | | | Not related n(%)E | | | Total |
|---|---|---|---|---|---|---|---|---|
| **System organ class** | **Preferred term** | **Mild** | **Moderate** | **Severe** | **Mild** | **Moderate** | **Severe** | **n(%)E** |
| Cardiac disorders | Chest pain | - | - | - | - | - | 1(3.8)1 | 1(3.8)1 |
| | Dizziness | 1(3.8)1 | - | - | - | - | - | 1(3.8)1 |
| | Syncope | - | - | - | 1(3.8)1 | - | - | 1(3.8)1 |
| Ear and labyrinth disorders | Ear pain | - | - | - | 1(3.8)1 | - | - | 1(3.8)1 |
| Gastrointestinal disorders | Abdominal pain | 1(3.8)1 | - | - | - | - | - | 1(3.8)1 |
| | Abdominal discomfort | - | - | - | - | 1(3.8)1 | - | 1(3.8)1 |
| Infections and infestations | Coronavirus infection | - | - | - | - | 5(19.2)5 | - | 5(19.2)5 |
| | Ear infection | - | - | - | - | 1(3.8)1 | - | 1(3.8)1 |
| | Gastroenteritis | - | - | - | 1(3.8)1 | - | - | 1(3.8)1 |
| | Genitourinary chlamydia infection | - | - | - | - | 1(3.8)1 | - | 1(3.8)1 |
| | Herpes simplex | - | - | - | 1(3.8)1 | - | - | 1(3.8)1 |
| | Infectious mononucleosis | - | - | - | - | 1(3.8)1 | - | 1(3.8)1 |
| | Lower respiratory tract infection | - | - | - | - | 1(3.8)1 | - | 1(3.8)1 |
| | Nasopharyngitis | - | - | - | 1(3.8)2 | 3(11.5)3 | - | 4(15.4)5 |
| | Periodontitis | - | - | - | - | 2(7.7)2 | - | 2(7.7)2 |
| | Sinusitis | - | - | - | - | 1(3.8)1 | - | 1(3.8)1 |
| | Tonsillitis | - | - | - | 1(3.8)1 | 1(3.8)1 | - | 2(7.7)2 |
| | Vaginal infection | - | - | - | 1(3.8)1 | - | - | 1(3.8)1 |
| | Vulvovaginal candidiasis | - | - | - | - | 2(7.7)2 | - | 2(7.7)2 |
| Injury, poisoning and procedural complications | Ankle fracture | - | - | - | - | 1(3.8)1 | - | 1(3.8)1 |
| | Hand fracture | - | - | - | - | 1(3.8)1 | - | 1(3.8)1 |
| | Joint injury | - | - | - | 1(3.8)1 | - | - | 1(3.8)1 |
| Investigations | Blood bilirubin increased | - | - | - | 1(3.8)1 | - | - | 1(3.8)1 |
| | Blood phosphorus increased | - | - | - | 3(11.5)3 | - | - | 3(11.5)3 |
| | Blood pressure increased | - | - | - | 1(3.8)1 | 1(3.8)1 | - | 2(7.7)2 |
| | Heart rate abnormal | 1(3.8)1 | - | - | 1(3.8)1 | - | - | 2(7.7)2 |
| | Hepatic enzyme increased | - | - | - | 2(7.7)2 | - | - | 2(7.7)2 |
| | Human papilloma virus tests positive | - | - | - | 3(11.5)3 | - | - | 3(11.5)3 |
| | Neutrophil count decreased | - | - | - | 2(7.7)2 | - | - | 2(7.7)2 |
| | Neutrophil count increased | - | - | - | 1(3.8)1 | - | - | 1(3.8)1 |
| | White blood cell counts increased | - | - | - | 1(3.8)1 | - | - | 1(3.8)1 |
| Metabolism and nutrition disorders | Hyperuricaemia | - | - | - | 1(3.8)1 | - | - | 1(3.8)1 |
| Musculoskeletal and connective tissue disorders | Costochondritis | - | - | - | - | 1(3.8)1 | - | 1(3.8)1 |
| Renal and urinary disorders | Urinary tract infection | - | - | - | - | 2(7.7)2 | 1(3.8)1 | 3(11.5)3 |
| Reproductive system and breast disorders | Dysmenorrhoea | - | - | - | 1(3.8)1 | - | - | 1(3.8)1 |
| | Intermenstrual bleeding | 1(3.8)2 | - | - | 1(3.8)1 | - | - | 2(7.7)3 |
| | Menstruation irregular | - | - | - | 3(11.5)3 | - | - | 3(11.5)3 |
| | Pelvic pain | 4(15.4)4 | - | - | 1(3.8)1 | 1(3.8)1 | - | 6(23.1)6 |
| | Vaginal discharge | 2(7.7)2 | - | - | - | 1(3.8)1 | - | 3(11.5)3 |
| | Vaginal odour | - | - | - | 1(3.8)1 | - | - | 1(3.8)1 |
| | Vulvovaginal dryness | 1(3.8)1 | - | - | - | - | - | 1(3.8)1 |
| Skin and subcutaneous tissue disorders | Acne | - | - | - | - | 1(3.8)1 | - | 1(3.8)1 |
| | Paraesthesia | - | - | - | 2(7.7)2 | - | - | 2(7.7)2 |
| | | | | | | | Total | 23(88.5)76 |

N=Participant Count; E=Event Count

The categories definite, probable, and possibly related are collapsed into the related category. The categories unlikely to be related and not related are collapsed into the not related category

**Supplementary Table 5**. Full list of adverse events (AEs) grouped by system organ class and whether the AEs were deemed related or unrelated to the intervention in the Active group (ITT population). All related AEs were classified as mild. N = participant count; E = event count. Participant count included individuals with at least one event; if they reported more than one AE, they were only counted once for the n. The % was calculated based on the number of participants in the group that reported at least one AE. Total events include all events reported regardless of whether a participant reported more than one.

| | | Related n(%)E | | | Not related n(%)E | | | Total |
|---|---|---|---|---|---|---|---|---|
| System organ class | Preferred term | Mild | Moderate | Severe | Mild | Moderate | Severe | n(%)E |
| Cardiac disorders | Dizziness | 1(12.5)1 | - | - | - | - | - | 1(12.5)1 |
| Gastrointestinal disorders | Abdominal pain upper | - | - | - | - | 1(12.5)1 | - | 1(12.5)1 |
| Infections and infestations | Coronavirus infection | - | - | - | 1(12.5)1 | 1(12.5)1 | - | 2(25.0)2 |
| | Gastroenteritis | - | - | - | - | 1(12.5)1 | - | 1(12.5)1 |
| | Lower respiratory tract infection | - | - | - | - | 1(12.5)1 | - | 1(12.5)1 |
| | Nasopharyngitis | - | - | - | - | 2(25.0)2 | - | 2(25.0)2 |
| | Oral herpes | - | - | - | - | 1(12.5)1 | - | 1(12.5)1 |
| Investigations | Blood phosphorus increased | - | - | - | 2(25.0)2 | - | - | 2(25.0)2 |
| | Blood pressure increased | - | - | - | 1(12.5)1 | 1(12.5)1 | - | 1(12.5)1 |
| | Neutrophil count decreased | - | - | - | 1(12.5)1 | 1(12.5)1 | - | 1(12.5)1 |
| | Neutrophil count increased | - | - | - | 1(12.5)1 | 1(12.5)1 | - | 1(12.5)1 |
| Renal and urinary disorders | Urinary tract infection | - | - | - | 1(12.5)1 | 1(12.5)1 | - | 1(12.5)1 |
| Reproductive system and breast disorders | Menstruation irregular | - | - | - | 1(12.5)1 | 1(12.5)1 | - | 2(25.0)2 |
| | Vaginal Discharge | - | - | - | 1(12.5)1 | - | - | 1(12.5)1 |
| Surgical and medical procedures | Tooth repair | - | - | - | 1(12.5)1 | - | - | 1(12.5)1 |
| | | | | | | | Total | 8(100.0)19 |

The categories definite, probable and possibly related are collapsed into the related category. The categories unlikely to be related and not related are collapsed into the not related category

**Supplementary Table 6**. Full list of adverse events (AEs) grouped by system organ class and whether the AEs were deemed related or unrelated to the intervention in the Placebo group (ITT population). All related AEs were classified as mild. N = participant count; E = event count. Participant count included individuals with at least one event; if they reported more than one AE, they were only counted once for the n. The % was calculated based on the number of participants in the group that reported at least one AE. Total events include all events reported regardless of whether a participant reported more than one.

| System organ class (SOC) | Active (n = 26) | | | Placebo (n=8) | | |
|---|---|---|---|---|---|---|
| Preferred term | Related | Not related | Total | Related | Not related | Total |
| | n(%)E | n(%)E | n(%)E | n(%)E | n(%)E | n(%)E |
| **Infections and infestations** | | | | | | |
| Ureaplasma infection | 7(27)7 | 0 | 7(27)7 | 1(13)1 | 0 | 1(13)1 |
| Staphylococcal infection | 0 | 1(4)1 | 1(4)1 | 0 | 0 | 0 |
| **Investigations** | | | | | | |
| Candida test positive | 2(8)2 | 0 | 2(8)2 | 1(13)1 | 1(13)1 | 2(25)2 |
| **Renal and urinary disorders** | | | | | | |
| Urinary tract infection | 1(4)1 | 0 | 1(4)1 | 0 | 0 | 0 |
| Asymptomatic bacteriuria | 0 | 1(4)1 | 1(4)1 | 0 | 0 | 0 |
| **Reproductive system and breast disorders** | | | | | | |
| Intermenstrual bleeding | 4(15)4 | 0 | 4(15)4 | 1(13)1 | 0 | 1(13)1 |
| Menstruation irregular | 3(12)3 | 0 | 3(12 3 | 1(13)1 | 1(13)1 | 2(25)2 |
| Coital bleeding | 2(8)2 | 0 | 2(8)2 | 0 | 0 | 0 |
| Vaginal discharge | 2(8)2 | 0 | 2(8)2 | 0 | 0 | 0 |
| Vaginal odor | 1(4)1 | 0 | 1(4)1 | 0 | 1(13)1 | 1(13)1 |
| Vaginal hemorrhage | 1(4)1 | 0 | 1(4)1 | 0 | 0 | 0 |
| **Totals** | 15(58)23 | 2(8)2 | 16(62)25 | 3(38)4 | 2(25)3 | 4(50)7 |

**Supplementary Table 7.** Full list of adverse events of special interest (AESIs) grouped by system organ class and whether the AESIs were deemed related or unrelated to the intervention in the Active and Placebo groups (ITT population). N = participant count; E = event count. Participant count included individuals with at least one event; if they reported more than one AE, they were only counted once for the n. The % was calculated based on the number of participants in the group that reported at least one AE. Total events include all events reported regardless of whether a participant reported more than one.
